# Data Resource Profile: Whole Blood DNA Methylation Resource in Generation Scotland (MeGS)

**DOI:** 10.1101/2024.04.30.24306314

**Authors:** Rosie M. Walker, Daniel L. McCartney, Kevin Carr, Michael Barber, Xueyi Shen, Archie Campbell, Elena Bernabeu, Emma Aitken, Angie Fawkes, Nicola Wrobel, Lee Murphy, Heather C. Whalley, David M. Howard, Mark J. Adams, Konrad Rawlik, Pau Navarro, Albert Tenesa, Cathie L Sudlow, David J Porteous, Riccardo Marioni, Andrew M. McIntosh, Kathryn L. Evans

**Affiliations:** Centre for Genomic and Experimental Medicine, Institute of Genetics and Cancer, University of Edinburgh, Edinburgh, UK; School of Psychology, University of Exeter, Exeter, UK; Medical Research Council Human Genetics Unit, Institute of Genetics and Cancer, University of Edinburgh, Edinburgh, UK; Division of Psychiatry, University of Edinburgh, Royal Edinburgh Hospital, Edinburgh, UK; Usher Institute of Population Health Sciences and Informatics, University of Edinburgh; Edinburgh Clinical Research Facility, University of Edinburgh, Western General Hospital, Edinburgh, UK; Institute of Psychiatry, Psychology and Neuroscience, King’s College London, UK; Baillie Gifford Pandemic Science Hub, University of Edinburgh, Edinburgh, UK; The Roslin Institute, Royal (Dick) School of Veterinary Studies, University of Edinburgh, UK; Health Data Research UK, London, UK

## Abstract

We have generated whole-blood DNA methylation profiles from 18,869 Generation Scotland Scottish Family Health Study (GS) participants, resulting in, at the time of writing, the largest single-cohort DNA methylation resource for basic biological and medical research: Methylation in Generation Scotland (MeGS). GS is a community- and family-based cohort, which recruited over 24,000 participants from Scotland between 2006 and 2011. Comprehensive phenotype information, including detailed data on cognitive function, personality traits, and mental health, is available for all participants. The majority (83%) have genome-wide SNP genotype data (Illumina HumanOmniExpressExome-8 array v1.0 and v1.2), and over 97% of GS participants have given consent for health record linkage and re-contact. At baseline, blood-based DNA methylation was characterised at ∼850,000 sites across four batches using the Illumina EPICv1 array. MeGS participants were aged between 17 and 99 years at the time of enrolment to GS. Blood-based DNA methylation EPICv1 array profiles collected at a follow-up appointment that took place 4.3-12.2 years (mean=7.1 years) after baseline are also available for 796 MeGS participants. Access to MeGS for researchers in the UK and international collaborators is via application to the GS Access Committee.

## Data resource basics

MeGS, which was initiated in 2016, was established to allow integration of whole blood DNA methylation data with the rich phenotypic, genetic, and electronic health record linkage already available for Generation Scotland (GS) a population- and family-based cohort (N=24,096 from 6,862 families) [1–3]. DNA methylation is an epigenetic modification influenced by both genetic and environmental factors, making it an attractive candidate for investigating mechanisms underlying complex traits and disease. In some circumstances, DNA methylation is associated with the expression of nearby or, less often, distal genes. There is clear evidence for associations between variation in DNA methylation and health-related behaviours, complex phenotypes, and disease outcomes [4–11]. MeGS, which is located in Edinburgh, UK was primarily funded through Wellcome Trust support.

GS was established through a multi-institutional collaboration, involving Scottish medical schools and the National Health Service [1,2]. It is an ideal cohort for the development of clinical and research biomarkers for use in disease prevention, detection, and monitoring, as whole blood is available from almost all participants, whilst linkage to health and prescription records allows for assessment of associations with both past and future health outcomes. Moreover, as participants have provided permission for re-contact, the potential to add additional longitudinal data points to MeGS exists, permitting studies of changes in methylation across the life course.

MeGS comprises blood-based DNA methylation profiles from 18,869 individuals making it, at the time of writing, the largest published single-cohort methylation resource in the world. For 796 of these participants, a second blood DNA methylation profile (the longitudinal MeGS sample) is available from an appointment that took place between 2015 and 2018 as part of a sub-study called “Stratifying Resilience and Depression Longitudinally” (STRADL). Key demographic information for MeGS is displayed in ***Table 1***. The MeGS cohort is broadly representative of the larger GS cohort, containing a higher proportion of females (58.8%), an average age of 47.12 years (SD 14.90 years), higher education levels and lower deprivation levels than the general population (***Table 1***). However, the participants for whom DNA methylation from a second time-point is available are older, have a greater average BMI, and are from less deprived localities than the baseline population (***Table 1***).

**Table 1:** Key demographic information.

|  | **GS Baseline (N=18,869)** | **STRADL Follow-up (N=796)** |
| --- | --- | --- |
| **Age (Years)** |  |  |
| Mean (SD) | 47.12 (14.90) | 56.49 (9.63) |
| Age Range | 17-99 | 20-82 |
| **Sex** |  |  |
| Males | 7771 (41.1%) | 342 (43.0%) |
| Females | 11094 (58.8%) | 454 (57.0%) |
| Not reported | 0 (0%) | 3 (0.4%) |
| **Education (Years)** |  |  |
| 0 Years (%) | 7 (0.04%) | 0 (0%) |
| 1-4 Years (%) | 50 (0.26%) | 4 (0.5%) |
| 5-9 Years (%) | 546 (2.9%) | 16 (2.0%) |
| 10-11 Years (%) | 4954 (26.25%) | 246 (30.9%) |
| 12-13 Years (%) | 3888 (20.6%) | 171 (21.5%) |
| 14-15 Years (%) | 2563 (13.58%) | 96 (12.1%) |
| 16-17 Years (%) | 3525 (18.68%) | 137 (17.2%) |
| 18-19 Years (%) | 1699 (9.0%) | 70 (8.8%) |
| 20-21 Years (%) | 431 (2.28%) | 15 (1.89%) |
| 22-23 Years (%) | 128 (0.68%) | 8 (1.0%) |
| 24+ Years (%) | 62 (0.33%) | 2 (0.25%) |
| Not Reported/Missing (%) | 1016 (5.4%) | 31 (3.89%) |
| **Deprivation (SIMD rank)** |  |  |
| Mean (SD) | 3898 (1848.55) | 4270 (1758.75) |
| **BMI (kg/m2)** |  |  |
| Mean (SD) | 26.67 (5.18) | 28.07 (5.58) |
| Range | 10.49-71.35 | 16.4-62.4 |
| **Smoking** |  |  |
| Ever (%) | 8630 (45.7%) | 326 (40.8%) |
| Never (%) | 9636 (51.1%) | 402 (50.3%) |
| Missing (%) | 603 (3.2%) | 72 (9%) |

Recruitment to GS is ongoing (“NextGenScot”, https://www.ed.ac.uk/generation-scotland) [12] with the aim of doubling the cohort size by recruiting additional members of existing families, new families, and lowering the minimum age of participation to 12 years. It is anticipated that saliva-based DNA methylation will be profiled in the newly recruited participants.

## Data collected

### Baseline Data and Sampling

The data and biological samples collected at the baseline clinic visit have been described in detail previously [2]. Participants completed a comprehensive pre-clinical questionnaire capturing information on various demographic variables, social characteristics, personal behaviours, cognitive and mental health and self-reported health data. Participants also attended a clinic, where physical and cognitive measurements were acquired, along with blood and urine samples.

The full range of phenotypic, clinical, and biochemical data available for GS (and, therefore, MeGS) participants has been described previously [1–3] and is searchable via the GS data dictionary (https://datashare.ed.ac.uk/handle/10283/2988). Genome-wide genotype information is available for the majority of MeGS participants (99.9%) [13]. When recruited between 2006-2011, GS participants provided blood or saliva samples (for biochemistry, and cryopreservation) and a urine sample, meaning it is possible to measure additional biomarkers in this cohort. As a condition of access by external researchers, any resource generated from GS must be returned to the study to be made available to the wider research community. This has resulted in the availability of cardiac and inflammatory biomarker data (NT-proBNP, GDF-15, cardiac troponins, and C-reactive protein) [14]. Furthermore, additional layers of omics data including mass spectrometry proteomics data (N = 15,818; N_MeGS_ = 14,671) and Nightingale metabolomics data (N = 2,907; N_MeGS_ = 2,745) have also been made available to GS through this route.

A subset of the MeGS cohort (44.4%) is also enrolled in the STRADL sub-study, in which questionnaires examining psychological resilience, coping style, threatening life experiences, and physical and mental health, were administered to 21,525 eligible GS participants, with 9,618 respondents (8,379 in MeGS) [15]. Of the 8,379 participants, 1,033 attended an in-person appointment where additional clinical and cognitive assessments were performed, blood, saliva, and hair samples collected, and neuroimaging performed [16].

Plasma levels of 4,058 proteins were measured using SOMAscan® V.4 technology in 839 MeGS participants from samples collected at the STRADL appointment [17]. Hormone levels were assayed from hair samples acquired as part of STRADL in 1,009 individuals, of whom 732 have methylation at both baseline and longitudinal time points. A subset of MeGS participants (N=4,233) also took part in the CovidLife surveys, which examined the effects of COVID-19 measures on health and well-being [18].

### Data linkage

Ninety-seven percent of GS participants consented to linkage to their NHS Scotland records. Linked datasets include hospital admissions from the Scottish Morbidity Record (SMR), dispensed prescription information, MIDAS dental data, and the Scottish Drug Misuse Database. SMR linkage includes general hospital admissions, maternity and neonatal data, psychiatric admissions, and diabetes and cancer registries. Mortality data is also available through linkage to the National Records of Scotland. New linkages, such as Scottish Medical Imaging (SMI), radiology reports, and retinal scans, are planned to continue following participants over time [3]. Linkage to primary care is also available for GS, albeit currently limited to a subset of 6,486 MeGS participants due to the constraints of data controller permission requirements.

## Sample processing

DNA methylation was profiled using the Infinium MethylationEPIC BeadChip v1 (Illumina Inc., CA), which measures methylation at over 850,000 sites across the human genome, covering 99% of RefSeq genes, and providing enhanced coverage of regulatory regions compared to previous Illumina methylation arrays. Peripheral blood samples were collected in EDTA tubes according to standard procedures and DNA extracted using Nucleon BACC3 extraction kits. Whole blood genomic DNA (500ng) was treated with sodium bisulphite using the EZ-96 DNA Methylation Kit (Zymo Research, Irvine, California), following the manufacturer’s instructions. DNA methylation was then measured using the MethylationEPIC BeadChip, according to the manufacturer’s instructions. Array scanning was performed using a HiScan scanner (Illumina Inc., San Diego, California) and an initial review of the data quality was carried out using GenomeStudio (version 2011.1).

DNA methylation was profiled in four waves between 2016 and 2021 (***Table 2***). For all waves, the raw intensity data (IDAT) files were read into R, using functions within minfi v.1.20.2 – 1.42.0 [19].

**Table 2:** Details of samples processed in each of the four methylation waves.

|  | **Wave 1** | **Wave 2** |  | **Wave 3** |  | **Wave 4** | **Combined** |
| --- | --- | --- | --- | --- | --- | --- | --- |
| **Date Prepared** | Q4 2016 – Q1 2017 | Q1 2017 – Q2 2017 |  | Q4 2018 – Q1 2019 |  | Q3 2021 | Q2 2023 |
| **Study Timepoint** | Baseline | Baseline | Longitudinal | Baseline | Longitudinal | Baseline | Baseline |
| **No. Samples (pre-QC)** | 5,200 | 465 | 520 | 4,597 | 373 | 9082 | 20,232 |
| **No. Samples (post-QC)** | 5,087 | 459 | 501 | 4,450 | 295 | 8,873 | 18,869 |
| **No. Probes (post-QC)** | 860,925 | 859,730 |  | 856,671 |  | 854,642 | 851,610 |

Quality control (QC) and normalisation was applied to each wave separately, at the time it was produced, to enable insights from these novel data as they emerged. Prior to commencing QC of the first wave, ten samples were removed as they were derived from saliva and were mistakenly submitted for whole blood methylation profiling. Three further samples were removed due to inaccurate self-report data (i.e., answering ‘Yes’ to all self-reported conditions). Another sample was removed as information from a separate study highlighted that they were likely to be XXY. For the first wave, QC was performed using shinyMethyl v1.10.0 [20]. First, technical outliers were removed based on visual inspection of a plot of the log median intensity of the methylated versus unmethylated signal for each array. ShinyMethyl’s control probe plots were then inspected to identify outliers. Next, samples for which the sex predicted from the methylation data (based on the difference between the median copy number intensity for the Y chromosome and the median copy number intensity for the X chromosome) did not match the participant’s self-reported sex were removed. Multi-dimensional scaling (MDS) plots were inspected for any additional sample outliers, but none were detected. The pfilter function within wateRmelon v.1.18.0 [21] was used to remove: (i) samples where ≥ 1% of CpGs had a detection *p*-value > 0.05; (ii) probes with a beadcount of < 3 in > 5% samples; and (iii) probes for which ≥ 0.5% of samples had a detection *p*-value > 0.05. Proportions of six white blood cell types (monocytes, granulocytes, CD 4+ T-cells, CD 8+ T-cells, B-cells, and natural killer cells) were estimated using minfi’s implementation of the Houseman algorithm [22] with Reinius et al.’s peripheral blood reference data [23].

The QC of the DNA methylation data produced in waves two to four was carried out using both meffil vs.1.1.0 and 1.1.2 [24] and shinyMethyl vs.1.14.0 and 1.30.0 [20]. Meffil was used to perform dye-bias and background correction using the “noob” method [25], and exclude: samples affected by a strong dye bias or issues affecting bisulphite conversion (using default thresholds); samples for which the median methylated signal intensity was more than three standard deviations lower than expected; and samples where the methylation-predicted sex deviated from self-reported sex. Deviations between methylation-predicted sex and self-reported sex were also assessed using shinyMethyl’s sex prediction function, which uses a different methodology to meffil. ShinyMethyl was additionally used to plot the output of all control probes to permit the detection of outliers by visual inspection. Following these sample removal steps, meffil was used to filter poor-performing samples and probes. Samples were removed if they had > 0.5% CpG sites with a detection *p*-value > 0.01. Once the poor-performing samples had been removed, meffil was re-run on the remaining dataset to identify poor-performing probes. These were defined as probes with a beadcount of < 3 in > 5% samples and/or > 1% samples with a detection *p*-value > 0.01. White blood cell proportions were estimated using meffil’s implementation of the Houseman algorithm [22] with Reinius et al.’s peripheral blood reference data [23].

Following QC, the data from Y chromosome probes were removed, and the data from each of the four waves were normalised using the dasen method from the R package wateRmelon v.2.2.0 [21], accounting for the EPIC arrays differing assay chemistries. Dasen involves adjusting the background difference between Type I assays (which interrogate methylated and unmethylated CpGs with separate probes) and Type II assays (which interrogate methylated and unmethylated CpGs with a single probe), by adding the offset between Type I and II probe intensities to Type I intensities. Between-array quantile normalisation is then performed for the methylated and unmethylated signal intensities separately, with Type I and Type II assays being normalised separately. The dasen-normalised beta-values were logit-transformed to methylation M-values, using the beta2M function in the R package lumi v.2.30.0 [26]. M-values can be denoted as log2((M + α)/(U + α)), where M corresponds to the methylated signal, U corresponds to the unmethylated signal, and α corresponds to a constant offset (usually 100) to regularise the Beta value when both M and U values are small. Following normalisation, X chromosome probes were excluded from the dataset together with probes that have been predicted to bind sub-optimally according to Zhou et al. (2017) [27] or McCartney et al. (2016) [28]. The normalised data (with and without the data from the X chromosome probes) were inspected by multidimensional scaling (MDS) plots to identify any remaining outlier samples. MDS plots were generated from subsets of the (i) 10,000 and (ii) 100,000 most variable probes, colour-coding by batch and sex. Visual inspection of these plots identified 40 male outliers in wave 3 who did not cluster with the other males. As a precaution, these samples were removed. For waves 2 and 3, the normalised datasets were subsequently separated into baseline and longitudinal sample sets. A jointly-normalised dataset is also available, whereby the four separately QC’d waves were combined and normalised using dasen. The dasen-normalised beta-values were converted to M-values as described above.

## Data Resource Use

The analysis of DNA methylation data can: provide novel insights into the mechanisms underlying diseases, health traits, and basic biological phenomena; identify biomarkers; and improve the prediction of future health outcomes. MeGS has contributed to research efforts across all these domains, resulting in 59 publications to date (Supplementary Table 1). We provide some examples below.

- **Methylome-wide association studies (MWASs):** MWASs have been carried out to identify associations between DNA methylation and a range of complex traits and diseases (Supplementary Table 1). As an illustrative example, we report here MWASs for two personality traits, neuroticism and extraversion, which have not been assessed previously in a large sample. MWASs were performed using *limma* v.3.54 [29]. Methylation levels in M-values (pre-corrected for relatedness using a genomic relationship matrix) at 752,721 CpG sites were included as the dependent variable and the and the independent variable was score on a personality trait (neuroticism: *n* = 18,788, extraversion: *n* = 18,783). The following model was fitted for each trait: *DNA methylation (corrected M-values) ∼ personality trait score + age + sex + methylation batch + estimated blood cell counts + methylation-derived smoking score + 20 methylation-based principal components*. A threshold of *p* < 3.6 × 10^−8^ was used to identify methylome-wide significant associations [30]. Fourteen CpG sites were identified as significantly associated with extraversion (**Figure *1A*** and ***Table 3***). No sites were significantly associated with neuroticism (**Figure *1B***).
- **Prediction of health outcomes and complex traits:** Barbu et al. (2022) [4] showed that a risk score calculated from methylation data explained 1.75% of the variance in major depressive disorder (MDD). McCartney et al. (2018) [9] derived DNA methylation predictors for 10 modifiable health and lifestyle factors and showed that a DNA methylation predictor of body mass index (BMI), when used in conjunction with a polygenic risk score, could explain approximately twice as much trait variance compared to the polygenic risk score alone. Cheng et al (2023) [31] have used MeGS to augment 10-year risk prediction of diabetes. Previous work on subsets of the MeGS baseline data has included the identification of methylation sites associated with: genetic risk factors for depression [32]; antidepressant treatment [33]; alcohol use disorder[34]; cognitive ability [7]; and risk factors for dementia [35–37]. MeGS has also contributed to collaborative meta-analyses to investigate DNA methylation associations with ageing [38], aggression [39], and chronic kidney disease [40]. In addition, the longitudinal dataset (i.e. methylation data acquired as part of STRADL) was used to generate signatures for 17 protein markers of brain health [41]. Finally, Chybowska et al. demonstrated the utility of methylation-derived EpiScores and composite measures of these scores to identify CVD risk, independent of traditional risk factors [42].
- **Validation of proxies for blood-based protein levels:** Gadd et al. related DNA methylation-based predictors of 109 protein levels (EpiScores) to incident health outcomes over 14 years. Using a subset of the MeGS cohort, they identified 137 EpiScore-disease associations, highlighting the potential of DNA methylation-based scores for disease prediction and risk stratification [17].
- **Epigenetic clocks:** The difference between a person’s actual age and their age predicted from their methylation data (age acceleration) provides a measure of biological ageing, which has been shown to be predictive of multiple health outcomes and all-cause mortality [43,44]. Using MeGS, we have demonstrated significant associations between age acceleration and several health-related traits, including BMI, smoking, socioeconomic status, and brain health [9,45]. In addition, we have shown that local CpG density affects the trajectory and variance of age-associated DNA methylation changes [46]. We performed genome-wide association study meta-analyses of four epigenetic clocks and discovered evidence for shared genetic loci associated with the Horvath clock and expression of lipid metabolism and immune function genes [38]. Finally, using the MeGS cohort, Bernabeu et al. derived a DNA methylation-based predictor of chronological age with a median absolute error of 1.7 years, outperforming existing predictors by between 1.8 and 6.4 years [47].
- **Improving understanding of basic biological mechanisms**: MeGS has contributed to a recent large-scale effort to map methylation quantitative trait loci (meQTL) [48]. This represents the most well-powered meQTL cataloguing effort to-date and resulted in the identification of > 270,000 independent meQTLs. The family-based nature of MeGS has facilitated studies into parent-of-origin effects on DNA methylation [49,50].
- **Methodological papers:** MeGS has been used to develop a Bayesian inference-based approach to analysis of methylation data [51], and has demonstrated the utility of Whatman FTA® cards for collecting and storing blood samples for DNA methylation profiling [52] MeGS has also contributed to a study assessing variability in DNA methylation-based predictors of age and BMI, when integrating multiple DNA methylation datasets. This study highlighted the importance of selecting an appropriate normalisation method for combining datasets and generating epigenetic signatures [53].

**Figure 1:**
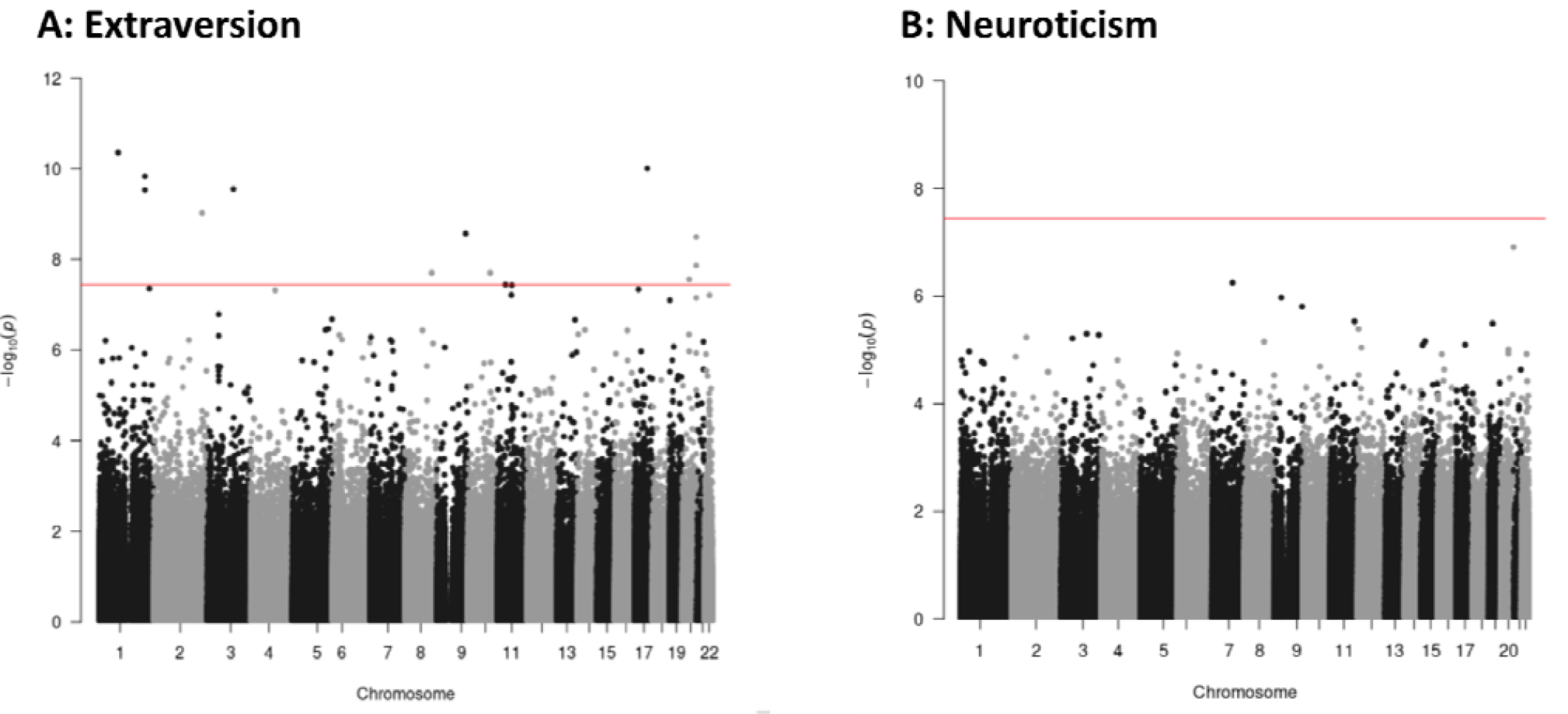
Manhattan plots of MWAS of Extraversion (A) and Neuroticism (B) Each point represents a CpG tested for association. -log_10_ p-values are presented along the Y-axis with chromosome and genomic position along the X-axis. The red line represents the epigenome-wide significant threshold of p=3.6 × 10^−8^.

## Strengths and Weaknesses

### Strengths

MeGS is derived from the Generation Scotland cohort, and is at the time of writing the largest single-population-based DNA methylation data resource. The use of the Illumina EPIC array, which almost doubles the coverage of its predecessor, the 450K array, is a further advantage over similar datasets [54]. Individuals in GS have been extensively phenotyped and the cohort is particularly well-suited for studies into mental health and cognitive phenotypes. In addition, record linkage to routine health datasets further increases the scope for investigations of both prevalent and incident diseases and traits. Finally, the ability to recontact participants for future studies permits longitudinal assessment.

**Table 3:** Differentially Methylated CpGs associated with Extraversion at p < 3.6×10^−8^.

| Probe ID | Effect | SE | P | Gene |
| --- | --- | --- | --- | --- |
| cg02947519 | $-9.14 \times 10^{-3}$ | $1.41 \times 10^{-3}$ | $1.05 \times 10^{-10}$ | - |
| cg10501210 | $6.20 \times 10^{-3}$ | $9.73 \times 10^{-4}$ | $1.96 \times 10^{-10}$ | - |
| cg15393490 | $8.93 \times 10^{-3}$ | $1.47 \times 10^{-3}$ | $1.18 \times 10^{-9}$ | - |
| cg05083539 | $8.24 \times 10^{-3}$ | $1.36 \times 10^{-3}$ | $1.46 \times 10^{-9}$ | *PKND; TMBIM1* |
| cg00329615 | $7.99 \times 10^{-3}$ | $1.30 \times 10^{-3}$ | $7.86 \times 10^{-10}$ | *IGSF11* |
| cg06690548 | $-1.45 \times 10^{-2}$ | $1.96 \times 10^{-3}$ | $1.82 \times 10^{-13}$ | *SLC7A11* |
| cg10585061 | $8.29 \times 10^{-3}$ | $1.50 \times 10^{-3}$ | $3.29 \times 10^{-8}$ | - |
| cg01182455 | $6.92 \times 10^{-3}$ | $1.20 \times 10^{-3}$ | $8.67 \times 10^{-9}$ | - |
| cg11376147 | $-7.51 \times 10^{-3}$ | $1.06 \times 10^{-3}$ | $1.20 \times 10^{-12}$ | *SLC43A1* |
| cg10861637 | $6.85 \times 10^{-3}$ | $1.12 \times 10^{-3}$ | $8.79 \times 10^{-10}$ | *CLTC* |
| cg11851174 | $8.67 \times 10^{-3}$ | $1.52 \times 10^{-3}$ | $1.34 \times 10^{-8}$ | *RAI1* |
| cg02231404 | $-8.97 \times 10^{-3}$ | $1.50 \times 10^{-3}$ | $2.45 \times 10^{-9}$ | *SOX18* |
| cg04881720 | $-9.88 \times 10^{-3}$ | $1.67 \times 10^{-3}$ | $3.66 \times 10^{-9}$ | *SOX18* |
| cg22138735 | $-8.49 \times 10^{-3}$ | $1.52 \times 10^{-3}$ | $2.60 \times 10^{-8}$ | *SOX18* |

### Weaknesses

The EPIC array characterises only ∼4% of the methylome, and cannot distinguish between DNA methylation and hydroxymethylation; however, it covers 99% of RefSeq genes, and has increased coverage of regulatory elements, such as enhancers, compared to previous Illumina methylation arrays. While blood is arguably not always the most mechanistically relevant tissue, the relative ease of obtaining peripheral blood samples makes it an appropriate tissue for development of biomarkers for prediction, monitoring disease course, and measuring the impact of treatment. In common with many other volunteer-based studies, GS has relatively small numbers of participants of non-white ancestry and more deprived backgrounds, and is not fully representative of the broader (Scottish) population.

## Data Resource Access

Researchers wishing to access the MeGS resource and wider Generation Scotland study data can do so by submitting an access application form to (contact person Dr D. McCartney). Access applications are subject to review through GS access processes, which ensure that all research using the resource aims to benefit the health and wellbeing of patients and the public. Approved projects are subject to a Data & Materials Transfer Agreement (DMTA) or commercial contract. Full information on the access procedure including application forms and DMTA templates is available at https://www.ed.ac.uk/generation-scotland/for-researchers/access. Data dictionaries describing the full GS resource are available online at https://datashare.ed.ac.uk/handle/10283/2988.

## Ethics approval

GS obtained ethical approval from the NHS Tayside Committee on Medical Research Ethics, on behalf of the National Health Service (reference: 05/S1401/89) and has Research Tissue Bank Status (reference: 20/ES/0021). All components of STRADL received formal, national ethical approval from the NHS Tayside committee on research ethics (reference 14/SS/0039).

## Author contributions

RMW, DLM, KC and KLE drafted the manuscript. EA, AF, NW, LM generated the raw DNA methylation data. RMW, DLM, AC, MJA and KLE were responsible for data management. RMW, DLM, KC, MB, XS, EB and KR performed quality control and data analysis with guidance from PN, AT, REM, AMM and KLE. DMH, HCW, DJP, AMM and KLE acquired funding to support the generation of the MeGS data resource within GS. CLS is the principal investigator of GS and acquired funding to support NextGenScot. All authors reviewed and approved the final manuscript.

## Supplementary data

Supplementary Table 1 – Summary of published studies that have used the MeGS resource to date.

## Funding

MeGS was primarily funded through Wellcome Trust support (reference 104036/Z/14/Z, 220857/Z/20/Z). Additional funding came from: a NARSAD Young Investigator Grant from the Brain & Behavior Research Foundation (Ref: 27404; awardee: David M Howard); a JMAS SIM fellowship from the Royal College of Physicians of Edinburgh (Awardee: Heather C Whalley); and a NARSAD Independent Investigator Award from the Brain & Behavior Research Foundation (Ref: 21956; awardee: Kathryn L Evans). The Chief Scientist Office of the Scottish Government and the Scottish Funding Council (HR03006) provided core support for Generation Scotland: Scottish Family Health Study, alongside a grant from the Scottish Government Health Department, Chief Scientist Office (Number CZD/16/6). “NextGenScot” is funded by the Wellcome Trust (ref 216767/Z/19/Z). PN is funded by BBSRC grant BBS/E/RL/230001A and acknowledges support from the MRC Human Genetics Unit program grant, U. MC_UU_00007/10, and grant MC_PC_U127592696. For the purpose of open access, the author has applied a Creative Commons Attribution (CC BY) licence to any Author Accepted Manuscript version arising from this submission.

## Acknowledgements

We are grateful to all the families who took part, the general practitioners and the Scottish School of Primary Care for their help in recruiting them, and the whole Generation Scotland team, which includes interviewers, computer and laboratory technicians, clerical workers, research scientists, volunteers, managers, receptionists, healthcare assistants and nurses. We thank Stewart Morris, Profs. Jonathan Seckl, Chris Haley, Joanna Wardlaw, Alison Murray, Caroline Hayward, Nick Hastie, Ian Deary and Stephen Lawrie for analytical inputs, study design, funding acquisition. We are also grateful to Robin Flaig (GS Chief Operations Officer) and her team, and the GS Scientific Steering Committee (Prof. Dame Anna F Dominiczak, Chair; Dr. Christian Cole, Profs. Katie Wilde, Cathie Sudlow, Heather Whalley, Riccardo Marioni, Zosia Miedzybrodzka, Sandosh Padmanabhan, Shantini Paranjothy, and Blair Smith).

## Conflict of interest

Daniel L. McCartney is a part time employee of Optima Partners Ltd. Lee Murphy has received speaker and consultancy fees from Illumina. Andrew M. McIntosh has received research support from Eli Lilly, Janssen, and the Sackler Foundation. Andrew M. McIntosh has also received speaker fees from Illumina and Janssen and consulting fees. Riccardo E. Marioni has received speaker fees from Illumina. Cathie L. Sudlow is Director of the British Heart Foundation Data Science Centre and Chief Scientist of the multifunder institute, Health Data Research UK.

